# A recurrent *SHANK3* frameshift variant in Autism Spectrum Disorder

**DOI:** 10.1101/2021.05.01.21256144

**Authors:** Livia O Loureiro, Jennifer L Howe, Miriam Reuter, Alana Iaboni, Kristina Calli, Delnaz Roshandel, Iva Pritisanac, Alan Moses, Julie D. Forman-Kay, Brett Trost, Mehdi Zarrei, Olivia Rennie, Lynette Lau, Christian R Marshall, Siddharth Srivastava, Brianna Godlewski, Elizabeth Buttermore, Mustafa Sahin, Dean Hartley, Thomas Frazier, Jacob Vorstman, Stelios Georgiades, Suzanne ME Lewis, Peter Szatmari, Lisa Bradley, Richard Delorme, Thomas Bourgeron, Evdokia Anagnostou, Stephen W. Scherer

**Affiliations:** Genetics and Genome Biology and The Centre for Applied Genomics, The Hospital for Sick Children, Toronto, ON, CAN; Canada’s Genomics Enterprise (CGEn), The Hospital for Sick Children, Toronto, ON, CAN; Holland Bloorview Kids Rehabilitation Hospital, Toronto, ON, CAN; Department of Medical Genetics, BC Children’s Hospital Research Institute, University of British Columbia, Vancouver, BC, CAN; Program in Molecular Medicine, The Hospital for Sick Children, Toronto, ON, CAN; Department of Cell & Systems Biology, University of Toronto, Toronto, ON, CAN; Department of Biochemistry, University of Toronto, Toronto, ON, CAN; Genome Diagnostics, Department of Paediatric Laboratory Medicine, The Hospital for Sick Children, Toronto, ON, CAN; Department of Laboratory Medicine and Pathobiology, University of Toronto, Toronto, ON, CAN; Department of Neurology, Rosamund Stone Zander Translational Neuroscience Center, Boston Children’s Hospital, Harvard Medical School, Boston, MA, USA; Autism Speaks, New York, NY, USA; Autism Speaks and Department of Psychology, John Carroll University, Cleveland, OH, USA; Department of Psychiatry, University of Toronto, Toronto, ON, CAN; Department of Psychiatry, The Hospital for Sick Children, Toronto, ON, CAN; Department of Psychiatry and Behavioural Neurosciences, McMaster University, Hamilton, ON, CAN; Centre for Addiction and Mental Health, Toronto, ON, CAN; Child and Adolescent Psychiatry Department, Robert Debré Hospital, Paris, France; Human Genetics and Cognitive Functions, Institut Pasteur, Paris, France; Department of Paediatrics, University of Toronto, Toronto, ON, CAN; Department of Molecular Genetics and the McLaughlin Centre, University of Toronto, Toronto, ON, Canada

## Abstract

Autism Spectrum Disorder (ASD) is genetically complex, but specific copy number variants (CNVs; e.g., 1q21.1, 16p11.2) and genes (e.g., *NRXN1, NLGN4*) have been identified as penetrant susceptibility factors, and all of these demonstrate pleiotropy. Many ASD-associated CNVs are, in fact, genomic disorder loci where flanking segmental duplications lead to recurrent deletion and duplication events of the same region in unrelated individuals, but these lesions are large and involve multiple genes. To identify opportunities to establish a more specific genotype and phenotype correlation in ASD, we searched genomic data, and the literature, for recurrent predicted damaging sequence-level variants affecting single genes. We identified 17 individuals from 15 unrelated families carrying a heterozygous guanine duplication (rs797044936; NM_033517.1; c.3679dup; p.Ala1227Glyfs*69) occurring within a string of 8 guanines (at genomic location [hg38]g.50,721,512dup) affecting *SHANK3*, a prototypical ASD gene (6/7,521 or 0.08% of ASD-affected individuals studied by whole genome sequencing carried the p.Ala1227Glyfs*69 variant). This variant, which is predicted to cause a frameshift leading to a premature stop codon truncating the C-terminal region of the corresponding protein, was not reproducibly found in any of the control groups we analyzed. All probands identified carried *de novo* mutations with the exception of five individuals in three families who inherited it through somatic mosaicism. This same heterozygous variant in published mouse models leads to an ASD-like phenotype. We scrutinized the phenotype of p.Ala1227Glyfs*69 carriers, and while everyone (16/16) formally tested for ASD carried a diagnosis, there was variable expression of core ASD features both within families and between families, underscoring the impact of as yet unknown modifiable factors affecting expressivity in autism.

---

Autism Spectrum Disorder (ASD) is a heterogeneous condition, both in clinical presentation and in terms of the underlying etiology. Individuals with ASD are increasingly being seen in clinical genetics^1,2^. More than 100 genetic disorders that can exhibit features of ASD (e.g. Fragile X, Phelan-McDermid syndromes, Rett)^3^ and dozens of rare susceptibility genes (e.g. *NLGN, NRXN, SHANK* family genes) and copy number variation (CNV) loci (e.g. 1q21.1 duplication,15q11-q13 duplication, 16p11.2 deletion), have been identified, which combined can facilitate a molecular diagnosis in ∼5–40% of ASD cases^4–7^. The likelihood of a genetic finding in ASD is dependent on the complexity of the phenotype (e.g. idiopathic or syndromic, with or without intellectual disability)^8,9^, the genomic technology used (e.g. microarrays, exome sequencing, genome sequencing, or combinations thereof)^10^, as well as the annotation pipeline and ‘gene lists’ used for interpretation^11,12^.

There are examples of how understanding the genetic subtypes of ASD can assist early identification enabling earlier behavioural intervention, and informing prognosis, medical management, and assessment of familial recurrence risk^13,14^. Moreover, genomic data promises to facilitate pharmacologic-intervention trials through stratification based on pathway profiles^15,16^. To support these applications, there is a growing interest in performing robust genetic analyses, often in families and in unique populations, linked to deep phenotyping ^17–19^.

The largest datasets available for genotype/phenotype correlations in ASD studies are based on CNV assessment since microarrays became the first-tier clinical diagnostic test^20,21^. The most relevant finding from this vast literature is that even for recurrent CNVs (i.e., genomic disorders) involved in ASD, which typically affect the same genes, there is variable expression of phenotypes relevant to the core features in autism, and other medical features^22–25^.

More recently, genotype and phenotype studies of sequence-level variation (single nucleotide variants, or SNV, and insertion/deletion, or indel, events) affecting individual genes are starting to reveal clinical correlations in ASD. For example, loss of function variants in the *SCN2A* sodium channel gene impairs glutamatergic neuronal excitability, leading to ASD and/or intellectual disability, while gain of function variants potentiate excitability leading to infantile-onset seizure phenotypes^26^. Different germline dominant-acting mutations in the phosphatase and tensin homolog (*PTEN*) gene found in ASD lead to an increased average head circumference in children^27^. Loss-of-function variants in the *CHD8* chromodomain helicase DNA-binding protein eight gene are also found in overgrowth and intellectual disability forms of ASD^28^. Despite some progress in resolving genotype-phenotype correlations, the vast genetic complexity and variable expressivity of genes involved in ASD continue to confound most predictive studies.

To minimize genetic complexity, here we searched our databases, other databases, and the literature for recurrent sequence-level damaging variants (*de novo* loss of function or missense variants predicted to be damaging based on the American College of Medical Genetics guidelines^29^) affecting the same site (genomic location) in the same gene in different families. In our most compelling finding, we identified a mutational ‘hotspot’ in a string of 8-Gs in exon 21 (p.Ala1227Glyfs*69) of the *SHANK3* gene that was present in 16 individuals from 14 unrelated families with ASD, as well as one individual with several autistic features and Phelan-McDermid Syndrome (but who was not tested for ASD). We assessed the intra- and inter-familial phenotypic variation (as well as all other genetic information) within these individuals and discuss the findings in the context of genotype-phenotype comparison including variable expression of ASD core symptom and related features.

To achieve the most comprehensive genomic representation (difficult to sequence exons, splice site boundaries) for variant detection, we initially examined the Autism Speaks MSSNG whole-genome sequencing (WGS) cohort (https://research.mss.ng/), with 11,359 samples, including 5,102 affected individuals and 3,567 with family data, typically belonging to trios, or quads (two parents and two affected children) for recurrent mutations. Secondly, we tested the Simon Simplex Collection (SSC) WGS collection (https://www.sfari.org/resource/simons-simplex-collection/), which comprises 9,205 samples, including 2,419 affected individuals and 2,393 with family data (typically two parents, one affected child, one unaffected child). Previous studies have extensively reported on the MSSNG^6,17,30,31^ and SSC^32,33^. Variant information and alignment files were downloaded from both databases. Probands from both cohorts met the criteria for ASD based on scores from standardized diagnostic criteria tools, typically the Autism Diagnostic Observation Schedule (ADOS)^34^ and the Autism Diagnostic Interview–Revised (ADI-R)^35^ and/or was supported by clinical criteria. Many individuals were also assessed with standardized measures of intelligence (I.Q.), including verbal and non-verbal ability, language, social behavior, adaptive functioning, and physical measurements^6,32,33^. Ethical review of these cohort studies was approved by institutional review boards.

From the genome sequences analyzed, our most interesting finding identified five probands in MSSNG (four males and one female) from four families and one proband in SSC (male) carrying a heterozygous guanine duplication in *SHANK3* (NCBI: NM_033517.1; ENSEMBL: ENST00000262795.5; c.3679 or c.3676 depending on the transcript) (Table 1; the reference sequence NM_033517.1 was selected as the appropriate transcript for this study as this was the reference sequence used in the original publication of this variant in Durand et al^37^). We also found other recurrent sequence-level *de novo* heterozygous damaging missense variants in the *PTEN, CAMK2A, SPTAN1, MECP2*, and *CSNK1E* genes, but in each of these instances, no more than two unrelated individuals were found in the combined MSSNG and SSC data (Supplementary Material; Table S1).

**Table 1.** Genome annotation of the *SHANK3* guanine duplication (rs797044936) considering different reference genomes, the position of the duplication in the guanine string, and the annotation tool. The guanine duplication in each carrier in the main text of the paper is referred to as p.Ala1227Glyfs*69.

| Reference Genome | Transcript Accession | Exon | Genomic Position | Coding change | Protein change | Annotation Tool |
| --- | --- | --- | --- | --- | --- | --- |
| Hg38 | NM_033517.1 | 21 | Chr22:50721512-50721513 | c.3679dup | p.(Ala1227Glyfs*69) | Alamut Visual v2.15.0<sup>#</sup> |
|  | NM_001080420.1<sup>##</sup> | 22 | Chr22:g.50721512dup | c.3727dup | p.(Ala1243Glyfs*69) | Alamut Visual v2.15.0 |
|  | ENST00000262795.5 | 24 | Chr22:g.50721512dup | c.3676dup | p.(Ala1226Glyfs*69) | Alamut Visual v2.15.0 |
|  | NM_001372044<br>(replaced NM_033517) | 22 | 22-50721503-50721504-T-TG | c.3855dupG | p.L1285fs | MSSNG |
| Hg19 | NM_033517.1 | 21 | Chr22:51159940-51159941 | c.3679dup | p.(Ala1227Glyfs*69) | Alamut Visual v2.15.0 |
|  | NM_001080420.1 | 22 | Chr22:g.51159940dup | c.3727dup | p.(Ala1243Glyfs*69) | Alamut Visual v2.15.0 |
|  | ENST00000262795.5 | 24 | Chr22:g.51159940dup | c.3676dup | p.(Ala1226Glyfs*69) | Alamut Visual v2.15.0 |
|  | NM_033517 | 21 | chr22:51159932-51159932-T-TG<br>chr22:51159933-51159933-G-GG | c.3630dup | p.L1210fs | VariCarta; GATK<br>VariCarta |
|  | ENST00000262795.3 | 22 | 22-5119932-T-TG | c.3719_3720<br>insG | p.Ala1243GlyfsTer6 | O'Roak et al.<sup>40</sup> |
|  | ENST00000262795.3 | 22 | 22-5119932-T-TG | c.3720dupG | p.L1240fs | Feliciano et al.<sup>41</sup> |

<sup>#</sup>Alamut Visual version 2.15 (SOPHiA GENETICS, Lausanne, Switzerland). This tool annotates the final G as duplicated and provides the coding and protein change as well as ClinVar entries and general population frequencies (Supplementary Figure S3).
<sup>##</sup> NM\_001080420.1 record has been removed from NCBI. This RefSeq was permanently suppressed because currently there is insufficient support for the transcript and the protein. Exon 11 was based on *ab initio* prediction and is not supported by transcript data.
At the time of submission, the most recent RefSeq NM\_01372044, which replaces and updates NM\_033517, was not available in the Alamut software.

The discovery of this recurrent guanine duplication variant in *SHANK3* was confirmed using Sanger sequencing (Figure 1). We then scanned the literature, including using Varicarta^36^ and found that this same guanine duplication was reported in 11 probands affected by ASD^4,37–42^, and one proband within the ASD borderline range, Phelan-McDermid syndrome, significantly delayed language, and speech and visual-motor deficits^38^. We carefully examined all genotypes and found that the proband reported by O’Roak et al.^40^ was the same individual in the SSC cohort (14470.p1); therefore, we removed this duplicate individual. Considering the new cases reported here and the cases reported in the literature, the p.Ala1227Glyfs*69 variant has been observed in a total of 17 cases from 15 families, identified using different genome-testing approaches (Table 2). Nearly all of these probands (16/17) were ascertained for ASD, although the general phenotype, as discussed below, varies somewhat among individuals (Table 3; Figure 2).

**Figure 1.**
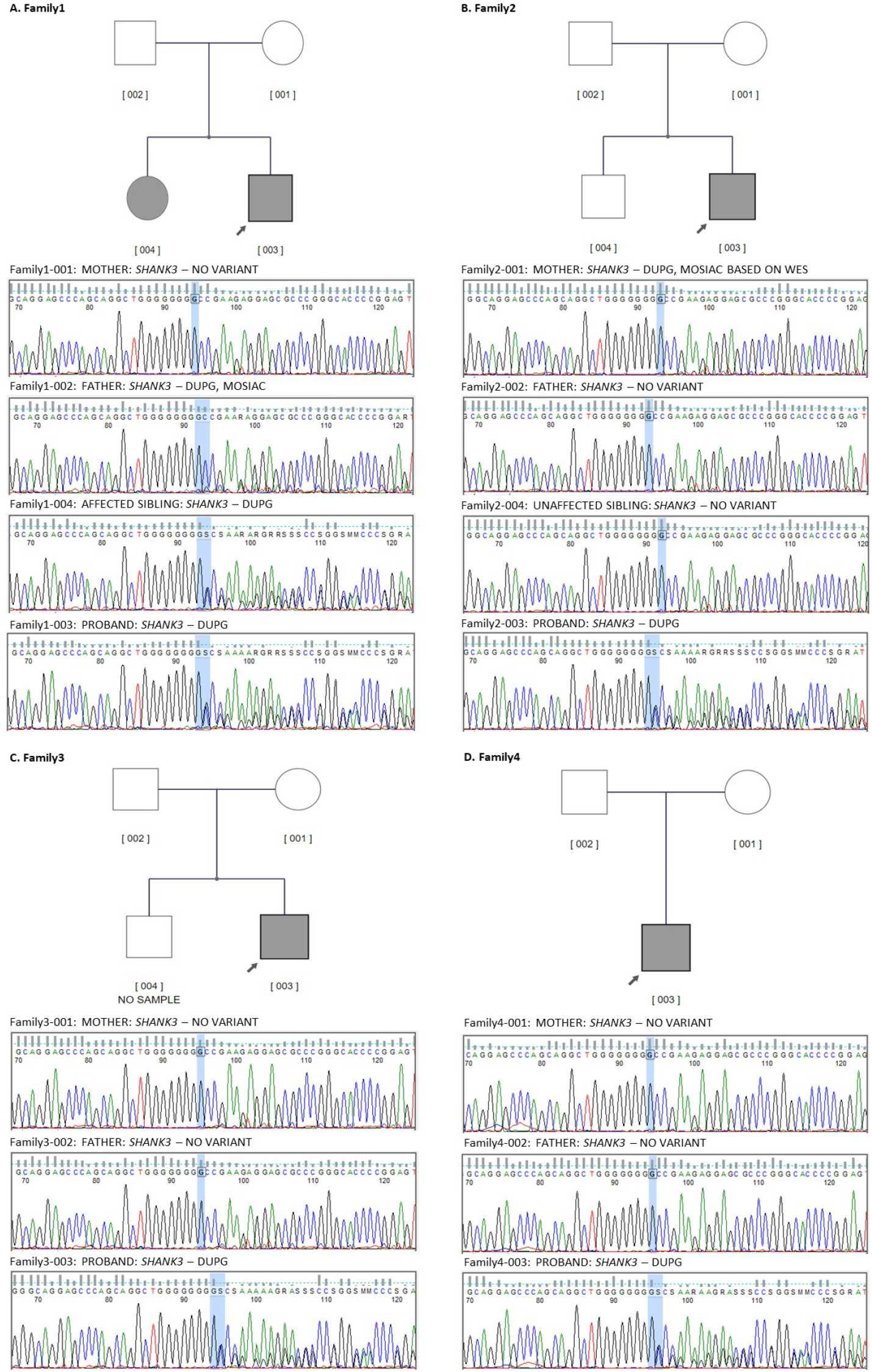
Pedigrees of MSSNG families reported for the first time in this study and their Sanger sequencing confirmation. A. Pedigree Family1; B. Pedigree Family2 (unaffected sibling was targeted Sanger sequenced but was not whole genome sequenced); C. Pedigree Family3 (unaffected sibling sample was not available); D. Pedigree Family4 (will be available in MSSNG DB7). Grey shapes indicate individuals with an ASD diagnosis and carry the *SHANK3* variant.

**Table 2.** ASD probands identified in MSSNG, SSC, and other publications containing the p.Ala1227Glyfs*69 *SHANK3* variant. Thicker lines box individuals from the same family. WGS – whole-genome sequencing, WES-whole-exome sequencing, PRS – polygenetic risk score, N/A – not available. Note that individual S7 described in De Rubeis^38^ has not been formally diagnosed with ASD but has reported autism-associated phenotypes. We also searched for p.Ala1227Glyfs*69 *SHANK3* variants in unpublished data from the SPARK cohort^41^. From 8,744 ASD-affected individuals for which sequencing data from both parents were available, the variant was detected in two male individuals, both *de novo*. The variant was also detected in 3 out of 13,156 ASD-affected individuals (two males and one female) for which parental sequences were not available and thus inheritance could not be determined. We mention this data just to demonstrate that the variant is found in other collections, as would be expected, and await the presentation of more detailed phenotype data from these participants.

| ID | SEX | GENOMIC (H.G.) | REFERENCE<br>SEQUENCE | CODING | PROTEIN | INHERITANCE | PUBLICATION/<br>COHORT | TECHNOLOGY | PRS<sup>1</sup> |
| --- | --- | --- | --- | --- | --- | --- | --- | --- | --- |
| Family1-003 | M | g.50721512dup (hg38) | NM_033517.1 | c.3679dup | p.A1227Gfs*69 | Paternal, mosaic | MSSNG – This paper | WGS | 3.639 |
| Family1-004 | F | g.50721512dup (hg38) | NM_033517.1 | c.3679dup | p.A1227Gfs*69 | Paternal, mosaic | MSSNG – This paper | WGS | 6.336 |
| Family2-003 | M | g.50721512dup (hg38) | NM_033517.1 | c.3679dup | p.A1227Gfs*69 | Maternal, mosaic | MSSNG – This paper | WGS | -1.167 |
| Family3-003 | M | g.50721512dup (hg38) | NM_033517.1 | c.3679dup | p.A1227Gfs*69 | *De novo* | MSSNG – This paper | WGS | 7.035 |
| Family4-003 | M | g.50721512dup (hg38) | NM_033517.1 | c.3679dup | p.A1227Gfs*69 | *De novo* | MSSNG-DB7 – This paper | WGS | 15.606 |
| ASD-2 pt1 | M |  | NM_033517.1 | c.3679dup |  | Maternal, mosaic | Durand et al.<sup>37</sup> | FISH and direct sequencing | Not estimated |
| ASD-2 pt2 | M |  | NM_033517.1 | c.3679dup |  | Maternal, mosaic | Durand et al.<sup>37</sup> | FISH and direct sequencing | Not estimated |
| S7 | F | g.51159940dupG (hg19) | NM_033517.1 | c.3679dupG | p.A1227Gfs*69 | Non-paternal | De Rubeis et al.<sup>38</sup> | WES | Not estimated |
| S8 | M | g.51159940dupG (hg19) | NM_033517.1 | c.3679dupG | p.A1227Gfs*69 | *De novo* | De Rubeis et al.<sup>38</sup> | WES | Not estimated |
| B1 | F | g.51159940dupG(hg19) | NM_033517.1 | c.3679dupG | p.A1227Gfs*69 | *De novo* | De Rubeis et al.<sup>38</sup> | WES | Not estimated |
| AU013503 | F | (hg19) | NM_033517.1 | c.3679dupG | p.Ala1227fs | *De novo* or father | Zhou et al.<sup>39</sup> | Target sequencing | Not estimated |
| AU035703 | F | (hg19) | NM_033517.1 | c.3679dupG | p.Ala1227fs | *De novo* | Zhou et al.<sup>39</sup> | Target sequencing | Not estimated |
| 14470.p1 | M | 22-51159932 -T-TG(hg19) | ENST00000262795.3 | c.3719_3720insG | p.Ala1243GlyfsTer6 | *De novo* | O'Roak et al.<sup>40</sup> | WES - O'Roak et al. 2014<br>WGS – This paper | 6.718 |
| ASD-685 | M |  | NM_033517.1 | c.3630dupG | p.L1210fs | *De novo* | Du et al.<sup>71</sup> | WES | Not estimated |
| G01-GEA-71-HI | F | 22:51159932:T:TG (hg19) |  |  |  | *De novo* | Satterstrom et al.<sup>4</sup> | WES | Not estimated |
| SP0051409 | F | 22- 51159932 -T-TG(hg19) | ENST00000262795.3 | c.3720dupG | p.L1240fs | *De novo* | Feliciano et al.<sup>41</sup> | WES | Not estimated |
| N/A | N/A | N/A | NM_033517.1 | c.3679dupG | p.A1227Gfs*69 | *De novo* | Farwell et al.<sup>42</sup> | WES | Not estimated |
| <sup>2</sup>ClinVar<br>SCV000850848<br>History of<br>Neurodevelopmental<br>Disorder | N/A | Chr22: g.50721512dup (hg38) | NM_033517.1 | c.3679dup | p.Ala1227fs | N/A | Ambry Genetics | Clinical testing | Not estimated |
| ClinVar<br>SCV000244220<br>Inborn genetic diseases | N/A | Chr22: g.50721512dup (hg38) | NM_033517.1 | c.3679dup | p.Ala1227fs | N/A | Ambry Genetics | Clinical testing | Not estimated |
| ClinVar SCV001149930<br>22q13.3 deletion syndrome | N/A | Chr22: g.50721512dup (hg38) | NM_033517.1 | c.3679dup | p.Ala1227fs | *De novo* | Institute of Human<br>Genetics, Klinikum rechts<br>der Isar | Clinical testing | Not estimated |
| ClinVar<br>SCV001149930<br>22q13.3 deletion syndrome | N/A | Chr22: g.50721512dup (hg38) | NM_033517.1 | c.3679dup | p.Ala1227fs | *De novo* | Institute for Genomic<br>Statistics and<br>Bioinformatics,<br>University Hospital Bonn | Clinical testing | Not estimated |
| ClinVar<br>SCV000329516<br>No condition provided | N/A | Chr22: g.50721512dup (hg38) | NM_033517.1 | c.3679dup | p.Ala1227fs | N/A | GeneDx | Clinical testing | Not estimated |
| ClinVar<br>SCV001468904<br>No condition provided | N/A | Chr22: g.50721512dup (hg38) | NM_033517.1 | c.3679dup | p.Ala1227fs | N/A | Laboratoire de<br>Génétique Moléculaire,<br>CHU Bordeaux | Clinical testing | Not estimated |

<sup>1</sup>For the PRS in addition to the main text only SNPs with minor allele frequency of > 0.05 in controls and high imputation quality (INFO > 0.9) were included. SNPs within the broad MHC region (Chr6:25–35MB) were excluded as well as all ambiguous SNPs to avoid potential strand conflicts. Only variants with good sequencing quality (filter = PASS) were included. Clumping was done with $r^2$ threshold and radius set at 0.1 and 500 kb, respectively. Subsequently, PRS was generated with p-value threshold of 0.1 weighing by the additive scale effect (log (OR)) of each variant and summing over the variants using PLINK 1.9. The scores were centered by the mean in whole population. <sup>2</sup>Variants submitted and catalogued by ClinVar (https://www.ncbi.nlm.nih.gov/clinvar/variation/208759/), Accession VCV000208759.8, provided from clinical testing and interpreted as pathogenic.

**Table 3.** Phenotype of ASD probands identified in MSSNG, SSC, and other publications containing the *SHANK3* p.Ala1227Glyfs*69 variant. Thicker lines box individuals from the same family. ID – intellectual disability; DD – developmental delay. ADHD - Attention deficit hyperactivity disorder, N/A – not available. Note that individual S7 described in De Rubeis et al.^38^ has not been diagnosed with ASD but has some autism-associated phenotypes. Graphical representation of these phenotypes is presented in Figure 2.

| ID | SEX | ASD | Dysmorphia | Intellectual disability / developmental delay | Other Medical comorbidities | Psychiatric comorbidity | Other Neurological comorbidities | Other Organ anomalies | Language/speech disorder |
| --- | --- | --- | --- | --- | --- | --- | --- | --- | --- |
| Family1-003 | M | Yes | Macrocephaly<br>Mandibular prognathism.<br>Malar flattening | Severe IDD |  | Bipolar disorder, Social Deficits |  | Conductive hearing loss | yes |
| Family1-004 | F | Yes | Macrocephaly<br>Asymmetric facial features (r; Mandibular prognathism; Prominent supraorbital ridge. Abnormal iris pigmentation. Hirsutism. Thick skin texture | Mild ID |  | Depression with psychotic tendencies, Self-injurious behaviour |  |  | yes |
| Family2-003 | M | Yes |  | Severe ID |  | ADHD | Epilepsy |  | severe |
| Family3-003 | M | Yes |  | Sever ID | Food sensitivities, G.I. distress, Eczema, Sleep disturbance | Anxiety | Epilepsy<br>Hypotonia, Hypermobility |  | Severe, with early language loss |
| Family4-003 | M | Yes |  | ID | Lactose intolerance | PICA (compulsive ingestion of inedible matter) |  |  | severe |
| ASD-2 pt1 | M | Yes |  | Severe ID |  |  |  |  | Severe |
| ASD-2 pt2 | M | Yes |  | Severe ID |  |  |  |  | severe |
| S7 | F |  |  | Mild ID |  |  | Hypotonia, Gait abnormality, Visuo-motor deficits, Self-stimulation | Coronary artery fistula | severe |
| S8 | M | Yes |  | Severe ID |  | Social Deficits | Hypotonia/Dysphagia, Abnormal gait |  | yes |
| B1 | F | Yes | Preauricular<br>skin tags<br>Finger and toe-<br>tapping | Mild ID | Scoliosis |  |  |  | yes |
| AU013503 | F | Yes |  | ? |  |  |  |  | yes |
| AU035703 | F | Yes |  | ? | Sleep<br>disturbance | hyperactivity | Abnormal gait |  | yes |
| 14470.p1 | M | Yes |  | ID |  |  | Epilepsy |  | Severe |
| ASD-685 | M | Yes |  | ID |  |  |  |  |  |
| G01-GEA-71-HI | F | Yes |  | ID |  |  |  |  |  |
| SP0051409 | F | Yes |  | ? |  |  |  |  |  |
| N/A | N/A | Yes |  | ? |  |  |  |  |  |

**Figure 2.**
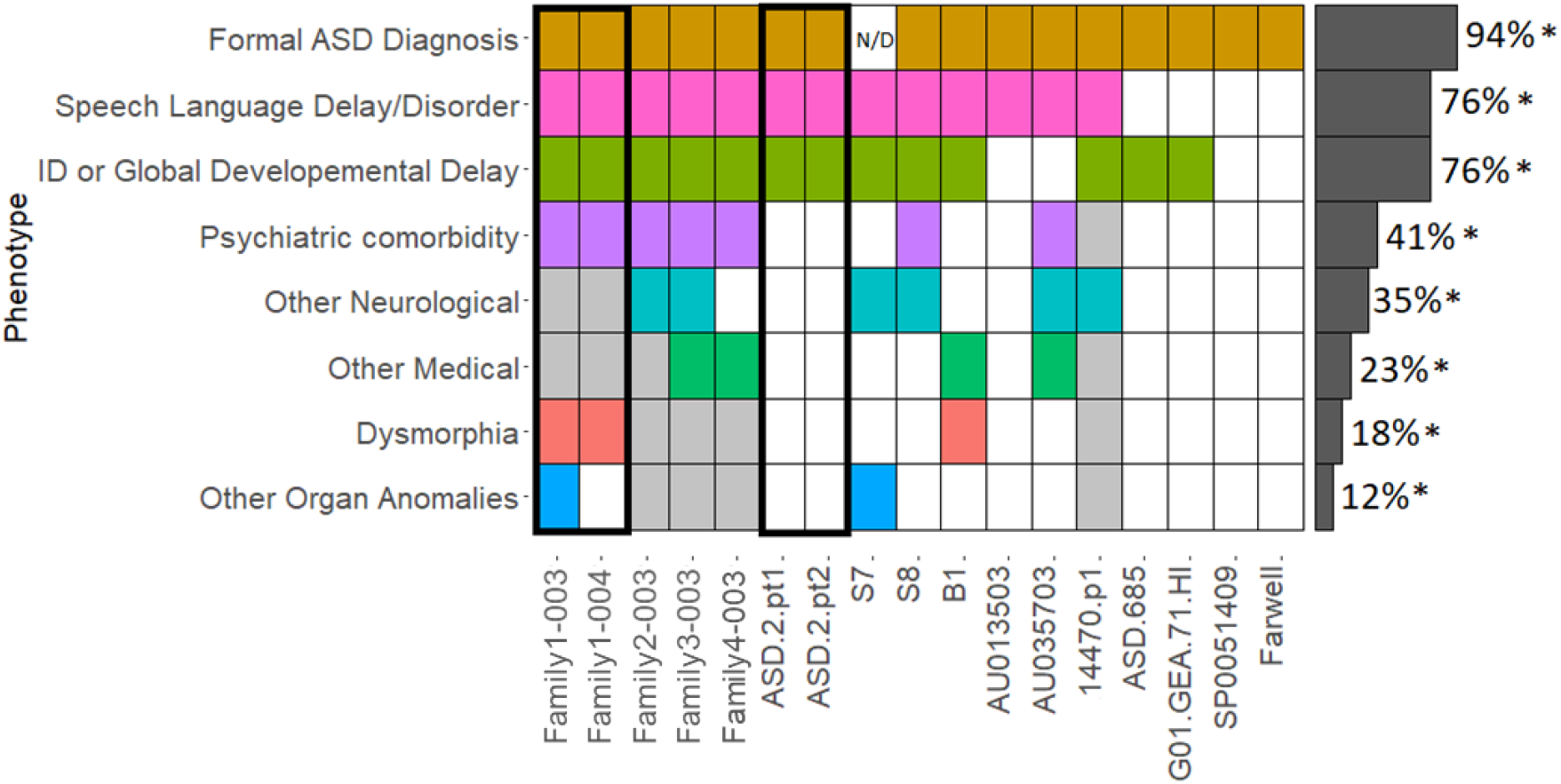
Phenotypic heterogeneity in individuals (X-axis) carrying the *SHANK3* p.Ala1227Glyfs*69 variant reported in the MSSNG^6,12^, SSC^32,32,33^, and in published papers^4,37-42,71^. Those individuals in the same family are grouped within the black boxes. Grey spaces indicate absence of the phenotype. White spaces indicate that the phenotype might have not been accessed in the proband. Phenotypic categories are described in Table 3. Individual S7 was not formally reported as being formally tested for ASD. * Caution is needed in the interpretation of these frequencies, since some phenotypes were not assessed for some individuals.

Given the high GC-density content of *SHANK3*, which can influence exon capture and sequencing^43^, we thought it was critical when assessing mutational frequency to confirm that there were no biases in read-coverage of the site of the target variant within exon 21 (Supplemental Material; Fig.1). Using whole exome sequences from 298 patients and 462 controls from our internal dataset, we ran the Agilent Sureselect Clinical research exome V1 for exome sequence analysis and show that the coverage around the G duplication region is at the anticipated 120x coverage (Supplemental Material; Fig. 1). This analysis also indicates that diagnostic exome sequencing will more than adequately capture and accurately genotype this position. WGS analysis of probands from MSSNG and SSC also confirm that exon 21 in *SHANK3* is uniformly covered.

All the probands identified in this study carried *de novo* variants with the exception of five individuals. One family with two brothers first reported in the initial *SHANK3* ASD-discovery paper^37^ inherited the variant from their mother, who was found to be mosaic. Two siblings within the MSSNG cohort (Family1-003 and Family1-004) inherited the variant from their father, who was also shown to be a mosaic (Table 2). In this latter case, the variant was only present in 8 of 50 reads in the father’s WGS data and was verified using a T.A. clone Kit (Invitrogen cat number 45-0046). Proband Family2-003 also seems to have inherited the variant from his mother by somatic mosaicism, in whom the variant was present in 1 of 32 reads of the WGS data. Exome sequencing analysis was also performed in this mother, with the variant being observed in 2 of 110 reads. To search for additional potential relevant somatic mutations^44^, we tested the original alignment files in both cohorts using DeNovoGear’s dng-call method for the *SHANK3* locus^45^ using 0.8 as a posterior probability of a *de novo* mutation (ppDNM), but we did not find any other candidates. Considering the families studied in MSSNG and SSC (our most trusted dataset) 6/7,521(0.08%) ASD-affected individuals carried the p.Ala1227Glyfs*69 variant in 5/6,681 (0.07%) of families. The Fisher’s exact test of the association between the frequency in heterozygous individuals in ASD cases and control population databases has a P-value of 0.029.

The *SHANK3* guanine duplication is located within a segment of 8-G’s on chromosome 22q13 at genomic location [hg38]g.50,721,505dup or g.50,721,512dup, depending on the position that this variant is annotated in the guanines (Table1; Figure 2). Some tools annotate the first G as the duplication, and others annotate it as the final G (Supplementary Material; Fig. 3). The sequencing technology might also affect the variant annotation, with Sanger sequencing conventionally adding the G duplication at the 3’ end of the gene, as the first point of amino acid change, and Next Generation Sequencing usually left aligning the variant. Independent of the position of the base insertion in the 8-Gs, the frameshift starting in exon 21 results in the new reading frame ending with a stop codon at position 69, causing a truncation lacking the C-terminal region (Figure 3).

**Figure 3.**
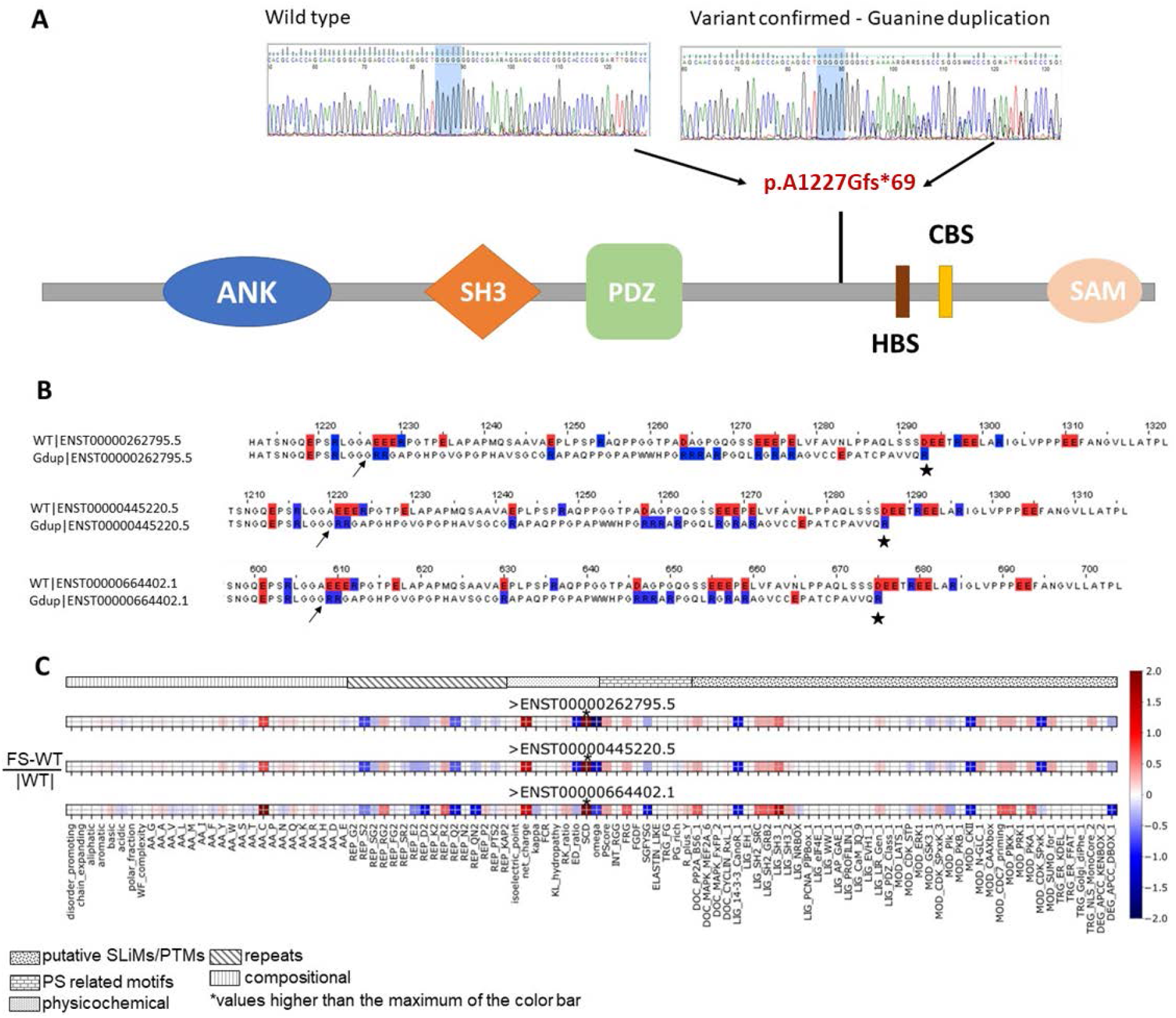
Impact of the *SHANK3* p.Ala1227Glyfs*69 variant on the protein **(A)** (top left) Guanine string containing 8 Gs found in non-affected individuals; (top right) Guanine string containing 9 Gs found in ASD affected individuals and parents with somatic mutations; (bottom) Location of the frequent guanine duplication in the *SHANK3* gene. ANK: ankyrin repeats; SH3: SRC homology 3 domain; PDZ: postsynaptic density 95/Discs large/zona occludens; HBS: homer binding site; CBS: cortactin binding site; SAM: sterile alpha motif domain; **(B)** Alignment of wild type protein sequences, for each of three highly expressed splice isoforms, to the protein sequence of the variant around the position of the mutation; (note, in this figure the first transcript presented is ENST00000262795.5 and the protein change for this is p.Ala1226Glyfs*69 as shown in Table 1) **(C)** Normalized impact of the variant for the three isoforms using FAIDR, a tool that identifies physical features and the presence of consensus protein recognition motifs in intrinsically disordered protein regions^56^. (*Note that SCD, sequence charge decoration, a measure of charge patterning associated with phase separation, has values significantly above 2: 5.4, 7.0, and 10.2 for the three isoforms.

Non-sense mutations and frameshifts in *SHANK3* can lead to reduced expression, and SHANK3-deficient neurons were found to have an altered phospho-proteome that may explain their decreased dendritic spine density^46^. However, *SHANK3* mRNA is still expressed in truncation mutant-containing induced pluripotent stem cells (iPSCs)^47^ and truncated SHANK3 proteins may have a dominant negative effects in neurons^48,49^. We therefore explored the consequences of p.Ala1227Glyfs*69 on the SHANK3 protein. We annotated the positions of amino acids to which the variant is mapped according to ENSEMBL and the UCSC genome browser. Using the DISOPRED3 predictor^50^ and the consensus of eight predictors from MobiDB-lite^51^, we identified where the mutation falls with respect to intrinsically disordered regions (IDRs) of the protein, which may influence protein folding and binding^52^. In both predictors, the position of interest was found to be embedded within a large IDR, which map to multiple isoforms (Figure 3B). Mutations that create frameshifts and stop codons in this region of *SHANK3*^37,43^ truncate two proline-rich binding sites for Homer and Cortactin (Figure 3A) and affect function, including altering neuronal morphology in cell-based experiments^47,48^. The SHANK3 protein serves as a scaffold to connect membrane receptors to the actin-cytoskeleton in the post-synaptic density (PSD), a protein-rich sub-compartment considered to be a biomolecular condensate formed by phase separation^53,54^ due to multivalent interactions^47^. In each of the isoforms, these truncations are expected to impair canonical PSD formation and stability.

The variant isoforms were also analyzed using Feature Analysis of Intrinsically Disordered Regions, a tool that identifies the presence of consensus protein recognition motifs in IDRs^55,56^ and using PScore^57^, predicts phase separation propensity via IDR planar pi-contacts (Figure 3C; Supplementary Material; Figure S2). A number of specific short linear interaction motifs were found to be altered. Of particular interest is the increase in SH3 domain class I-binding motifs, given that *SHANK3* is known to interact with numerous SH3 domains. The variants significantly increase the number of arginine-glycine and arginine-arginine dipeptide instances, which are associated with mRNA binding and phase separation, and increase the cysteine content of the sequence. A reduction in SHANK3 protein due to the frameshift (e.g., through non-sense mediated decay; discussed below) could also affect the phase separation of the PSD, which is known to be concentration dependent^58^.

The p.Ala1227Glyfs*69 variant is classified in ClinVar as “Pathogenic for ASD, NDD and others” and is exceptionally rare or absent in control populations (ClinVar; https://www.ncbi.nlm.nih.gov/clinvar/variation/208759/). In the gnomAD v2.1.1 dataset^59^, which uses the hg37 as reference genome, it has an allele frequency of 16/160,994 alleles = 0.00009938 (0.0099%). In ALFA^60^, this variant is also reported in 0.02% of control Europeans samples. However, in gnomAD v3, 1000 Genomes Project (that uses hg38 as a reference genome), TOPMed^61^, two unpublished paediatric controls from our group (INOVA and CHILD), the Personal Genome Project Canada^62^ and Medical Genome Reference Bank^63^ this variant is not present. In combination, this suggests that the presence of the variant in gnomAD v2.1.1 and ALFA might be due to low quality sequencing with the preliminary description being corrected in gnomAD v3. It is also noteworthy that ∼1/100 people will have ASD, so it would be expected to find p.Ala1227Glyfs*69 variant carriers in control populations. Based on our findings described here they would likely have ASD, but additional studies will be required to further assess this.

We have analyzed the genomic conservation of this variant with GERP^64^, UCSC PhyloP, and phastCons for primates, placental mammals, and 100 vertebrates^65^. GERP identifies constrained elements in multiple alignments by quantifying substitution deficits. These deficits represent substitutions that would have occurred if the element were neutral DNA but did not occur because the element has been under functional constraint. The p.Ala1227Glyfs*69 variant has a GERP score of 5.2 (p = 0), suggestive of having a large deleterious effect^66^. The PhyloP score was 0.6 for primates, 1.35 for mammals, and 2.13 considering 100 vertebrates, suggesting high evolutionary conservation. The PhastCon scores were also higher than 0.98 for primates, mammals, and vertebrates, which indicates a strong negative selection on this variant.

In all 16 p.Ala1227Glyfs*69 carriers evaluated for ASD, ASD was confirmed, and the majority of patients described are reported to have intellectual disability defined as an IQ score below 70 and impairments in adaptive functioning, although the spectrum of severity is wide (Table 3; Figure 2). Four individuals were ascertained for Phelan-McDermid Syndrome, with three of these being of the 16 receiving a formal ASD diagnosis and one never being assessed for autism. Language deficits are also prevalent and often severe. We were cautious about making claims on other associated conditions as they have not been universally and systematically ascertained. However, hypotonia and gait abnormalities are common, also consistent with animal model data^67^. Seizures were reported in 3/17 participants. Other neurodevelopmental concerns include ADHD, anxiety, and mood disorders. Gastrointestinal distress and sleep dysfunction were also reported. Lastly, both dysmorphia and other organ anomalies were reported (conductive hearing loss- and coronary artery fistula). Within pairs of siblings sharing a variant, there is a similarity of phenotype, with some variability in the severity of the intellectual disability.

Different *de novo* mutations in *SHANK3* have also been associated with other developmental/neuropsychiatric disorders and genetic syndromes such as schizophrenia^48,68^ and Phelan-McDermid Syndrome (PMS)^69^. The majority of children diagnosed with PMS also have ASD, and both conditions are often associated with intellectual and language delay, hypotonia, seizures, and sleep disorders, although children with PMS also often have other organ involvement. We also assessed other potential clinically relevant variants found in the p.Ala1227Glyfs*69 carriers to determine if they may contribute to varying phenotypic presentation either from the MSSNG and SSC probands or from those patients described in the literature^37,38,69–71^, but none were identified.

To evaluate if common genetic variants may be contributing to the ASD phenotype in the p.Ala1227Glyfs*69 *SHANK3* variant carriers, we calculated their ASD polygenic risk score (PRS). PRS was calculated for all individuals from European ancestry in MSSNG (db6) and SSC merged with 1000 Genomes European population using GWAS summary statistics derived from the iPSYCH Autism project including 13,076 cases and 22,664 controls from Denmark^72^. This included probands Family1-003, Family1-004, Family2-003, Family3-003, and 14470.p1. A total of 25,837 SNPs were included in PRS calculation. Since, the proband Family4-003 was part of a later version of the MSSNG cohort (db7), he was not included in the initial PRS calculation. This individual’s PRS was calculated separately with his parents (Family4-001 and Family4-002) using the same 25,837 SNPs included in PRS calculations for the others and centered by the mean in whole MSSNG/SSC/1000 Genomes European population. However, of 25,837 SNPs, 1,496 were missing due to sample quality in this family, and caution is needed in comparison with the other subjects. PRS in the probands analyzed in this study varied between -1.167 to 15.606 (Table 2), showing no clear pattern between the presence of the clinically significant *SHANK3* variant and the polygenetic risk of common variants. PRS in all subjects with autism in MSSNG and SSC ranges between -18.580 and 20.626.

Two independently-created murine models with an insertion of a guanine nucleotide into the analogous mouse base pair position, which we refer to here as Shank3 InsG3680, have also demonstrated changes in cellular, circuit and behavioural phenotypes^67,73,74^ (Supplementary Material; Table S2). Specifically, these Shank3InsG3680 mouse models demonstrated changes to baseline neurotransmission and/or impairments in long-term depression (LTD) and long-term potentiation (LTP), the synaptic basis of learning and memory. Overall homozygous Shank3InsG3680 +/+ mice exhibited more significant changes than heterozygous Shank3InsG3680 mice, suggesting that functioning of one normal Shank3 copy maybe sufficient to support some of its function.

Regional differences in synaptic deficits and synaptic composition were observed, and the extent of the impact may have been modulated by other Shank family genes. In the adult hippocampus, expression of the reversible Shank3InsG3680 variant cassette^67^ produced a truncated Shank3 protein and loss of the major high molecular weight isoforms at the synapse. This was associated with impaired hippocampal mGluR dependent LTD, intact LTP and changes to baseline NMDA receptor (NMDAR) mediated synaptic function. In the striatum, Zhou et al.^73^ showed a significant decrease of levels of Shank3 mRNA in the Shank3InsG3680 strain compared with the wild type, suggesting a reduced level of mRNA through non-sense-mediated decay. This finding suggests that the InsG3680 variant results in a near complete loss of SHANK3 protein, concomitant with synaptic transmission deficits in juvenile and adult homozygous mutant Shank3InsG3680 (+/+) mice. Post translational modifications, modulated by nitric oxide, were also found in both young and adult Shank3InsG3680 +/+mice^74^.

In assessments of general cognitive function, Shank3InsG3680 +/+ mice showed mild spatial learning impairments in the Morris Water Maze task and motor learning deficits in the accelerating rotarod task, while heterozygous mice did not^67^. ASD associated behaviors in these two models also showed mixed outcomes in both social interaction impairments and repetitive behaviors that, similar to human assessments, may be dependent on age and gender. Speed et al.^67^ reported statistically different effects in some of their assessments comparing between male and female adult mice. This group did not observe social interaction deficits in the three-chamber task with mixed sex adult mutant mice, nor did they observe repetitive behaviors, but instead suggested aversion to novel objects. However, in large all male cohorts, Zhou et al.^73^ showed deficits in social behaviors in both juvenile and adult mice. In addition, in adults there was increased anxiety, repetitive grooming behaviors and sensory processing differences^73^. On balance, the mouse data seems to generally recapitulate the learning impairments and behavioural differences seen in patients with the p.Ala1227Glyfs*69 *SHANK3* variant.

Highly penetrant alleles such as p.Ala1227Glyfs*69 in neurodevelopmental disorders are under severe negative selection and are constantly being removed from the population^75^. However, recurrent mutations are always being added to the gene pool and while typically occurring randomly, the intrinsic^76^ and extrinsic characteristics^77^ may also have an influence^78^. Experimental investigations have shown that guanine bases can be targets for oxidative damage in DNA, while mutability in other bases is more variable^79^. Moreover, the locus under study is within 8 guanines, which constitutes a homopolymer run (HR). HRs are sequences with six or more identical nucleotides and are associated with >10-fold enrichment of mutation compared to the genomic average^80^. It is noteworthy that there are three other G homopolymer runs in *SHANK3*, but no recurrent variants were found at these sites.

The CpG content of DNA has also been shown to influence the mutation rate in non-CpG-containing sequences, suggesting that intrinsic properties of DNA sequences may be more important than the chromosomal environment in determining mutation rates and genome integrity. Evidence indicates that because of the propensity for methyl-CpGs to deaminate and produce mismatches, it is plausible that error-prone repair mechanisms may have a role in hypermutability. CpG methylation might also have epigenetic effects by promoting chromatin states that make DNA more susceptible to mutations^81^.

In summary, our data indicate that 16/16 carriers (from 14 independent families) of the p.Ala1227Glyfs*69 variant affecting *SHANK3* who have been formally tested carry a diagnosis of ASD. Our analysis did not find any other obvious rare or common genetic variants, or combinations thereof, in the genomes of these families that seem to contribute to core ASD phenotype in these individuals. Given the nature of neurobehavioral complexity, perhaps not surprisingly, there is phenotypic heterogeneity exhibited amongst p.Ala1227Glyfs*69 carriers, which is a hallmark of autism^82,83^, as well as other related brain disorders that may share overlapping clinical features and contributory susceptibility genes^84,85^. Although exceedingly rare (0.075% frequency in the ASD families studied by WGS), the finding that this p.Ala1227Glyfs*69 variant in *SHANK3* is, so far, concordant with an ASD, and that it will surely continue to sporadically re-occur in the population, has important implications for genetic counselling. It will also be important to continue to search for the p.Ala1227Glyfs*69 variant in *SHANK3* to see if it confers risk in other disorders. Defining a specific mutational mechanism underlying an ASD outcome, may also focus strategies for the development of therapeutic interventions.

## Data Availability

Access to the MSSNG and SSC data can be obtained by completing data access agreements (https://research.mss.ng and https://www.sfari.org/resource/sfari-base, respectively), as was done for this study. These two well-established and stable whole genome sequence and phenotype resources are utilized by approved investigators worldwide. The 1000G genome-sequencing data are publicly available via Amazon Web Services (https://docs.opendata.aws/1000genomes/readme.html). Access to other data through other publications or resources is described in the main text.

https://www.sfari.org/resource/simons-simplex-collection/

## Acknowledgements

We thank the families for participation, The Centre for Applied Genomics and Verily Life Sciences for their analytical and technical support, as well as staff at Autism Speaks for organizational and fundraising support. We thank Jonathon Ditlev for insightful discussion. This work was funded by Autism Speaks, Autism Speaks Canada, the University of Toronto McLaughlin Centre, the Canada Foundation for Innovation, the Canadian Institutes of Health Research (CIHR), Genome Canada/Ontario Genomics Institute, the Government of Ontario, Brain Canada, Ontario Brain Institute Province of Ontario Neurodevelopmental Disorders (POND), and The Hospital for Sick Children Foundation. L.O.L holds Lap-Chee Tsui Postdoctoral Fellowship from The Hospital for Sick Children. S.W.S holds the Endowed Chair in Genome Sciences at the Hospital for Sick Children and University of Toronto.

