## Supplemental Material for "A recurrent *SHANK3* frameshift variant in Autism Spectrum Disorder"

**Supplementary Material**

Table S1 – Recurrent *de novo* damaging missense variants found in ASD probands in the MSSNG and SSC cohorts. Variants frequencies are less than 0.001 in gnomAD and 1000g.

| Gene | CHR | Start | End | Reference bases | Alternate bases | Protein Change | Sample ID | Sex |
|---|---|---|---|---|---|---|---|---|
| *PTEN* | chr10 | 87957940 | 87957941 | T | G | p.F241L | Family5-003 | F |
| | | | | | | | Family6-003 | M |
| *CSNK1E* | chr22 | 38300756 | 38300757 | G | A | p.R178C | Family7-003 | F |
| | | | | | | | Family8-003 | F |
| *CAMK2A* | chr5 | 150251807 | 150251808 | G | A | p.P212L | Family9-003 | M |
| | | | | | | | Family10-003 | M |
| *SPTAN1* | chr9 | 128632636 | 128632637 | T | C | c.T7142C | Family11-003 | M |
| | | | | | | | Family12-003 | F |
| *MECP2* | chrX | 154030911 | 154030912 | G | A | p.R213C | Family13-003 | F |
| | | | | | | | Family14-003 | F |

Table S2- Comparison between the phenotypes observed in two mouse models engineered to carry the murine equivalent of the *SHANK3* p.Ala1227Glyfs*69 variant found in humans. Phenotypes in humans are described in Table2.

| Phenotype | Mouse - *Shank3G* exon 21<br>Speed et al.[67] | Mouse - InsG3680<br>Zhou et al.[73] | Humans (Table 2) |
|---|---|---|---|
| Intellectual disability | x | x | x |
| Impaired motor coordination | x | x | x |
| Altered response to novelty | X (object) | x | |
| Anxiety-like behaviour | | x | x |
| Social interaction deficits | | x | x |
| Repetitive/compulsive behaviour | | x | |

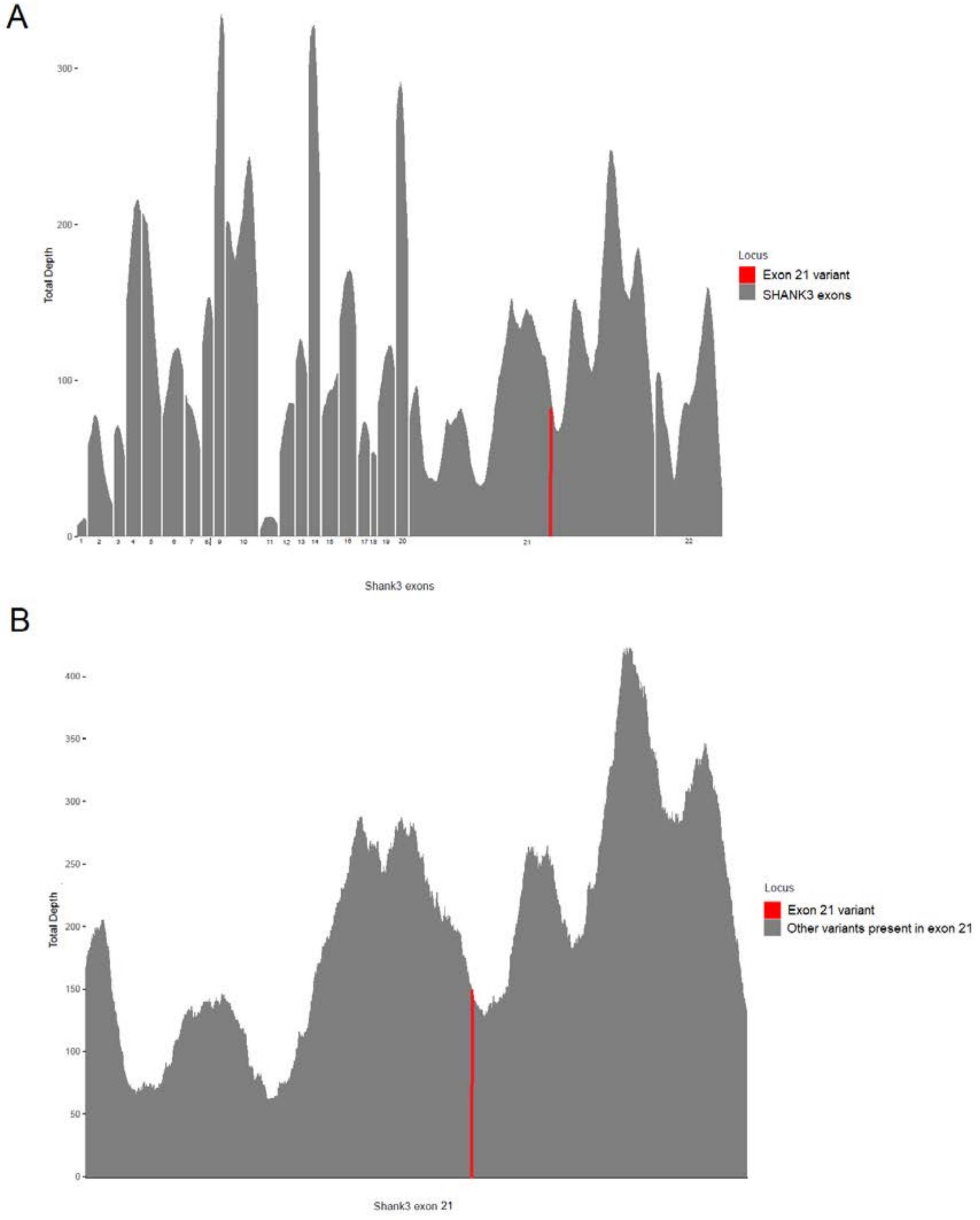


Figure S1 - A – *SHANK3* exon coverage per base pair calculated for exon 1 to 22 in 698 individuals. B – *SHANK3* exon 21 coverage calculated for 462 individuals. Red line indicates the position of the guanine duplication described in this study. Whole Genome sequencing also indicates high coverage sequencing across *SHANK3*, including in exon 21.

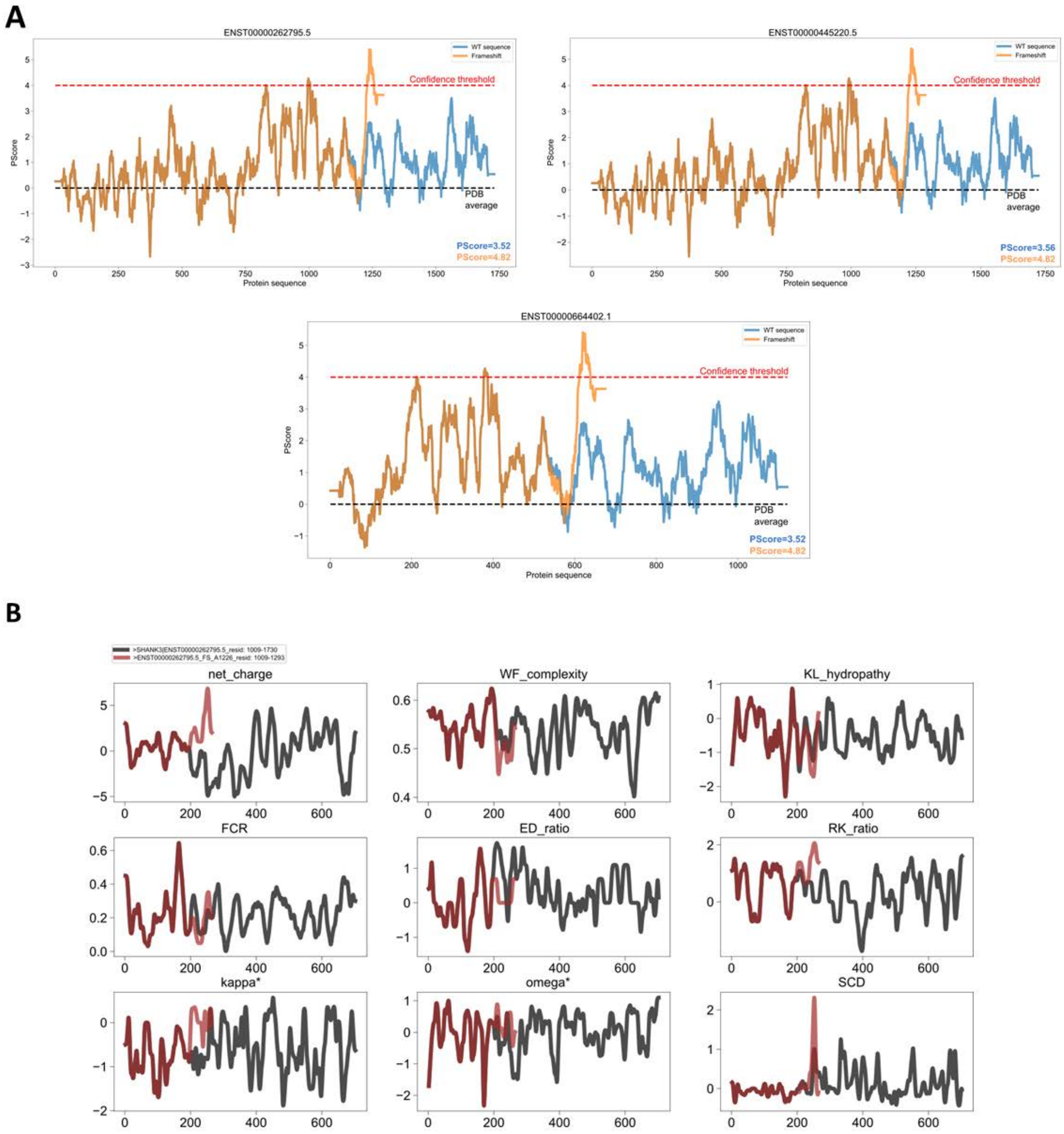


Figure S2. A- Sequence profiles of PScore[57], a predictor of phase separation via planar pi-pi interactions in intrinsically disordered proteins, for each of the three isoforms and the corresponding variant; B- Sequence profiles from FAIDR[56] for the isoform and the corresponding variant for the isoform ENST00000262795.5, highlighting different physicochemical features known to be associated with phase separation, including ratios of arginine to lysine (RK ratio) and two measures of charge patterning (kappa and sequence charge decoration, SCD). See Zarin et al.[55] for more details on these features.

***Warning:*** *This report is based on knowledge and data that are not firmly established. Consequently, medical decisions must not be made on the basis of this report.*

**SHANK3 (SH3 and multiple ankyrin repeat domains 3) Variation**

Duplication (1 bp) in exon 24.
This variation creates a frame shift starting at codon Ala1226. The new reading frame ends in a STOP codon at position 69.

**This variant is known to ClinVar** (February-2021): RCV000004730.7 (Pathogenic** - 22q13.3 deletion syndrome), RCV000190779.1 (Pathogenic* - Inborn genetic diseases), RCV000366708.2 (Pathogenic** - not provided), RCV000754675.1 (Pathogenic* - Autism spectrum disorder), RCV000719974.1 (Pathogenic* - History of neurodevelopmental disorder).

**This variant is known to dbSNP** (151): rs797044936 (validated dbSNP entry - Clinical significance: CLIN_pathogenic).

**This variant is known to ESP** (ESP6500SIV2): Eur. Am.: TGG=1.30% - Afr. Am.: TGG=1.01%

**This variant is known to gnomAD** (2.1) <Exomes>: ALL:0.0099% - AMR:0.0079% - ASJ:0.012% - SAS:0.0084% - NFE:0.014% - FIN:0.013% (**Filter**: RF)

HGVS Nomenclature (v15.11)

---

cDNA Level: **ENST00000262795.5:c.3676dup**
gDNA Level: **Chr22(GRCh38):g.50721512dup**
Protein Level: **p.(Ala1226Glyfs*69)**

---

©Interactive Biosoftware - Created by Alamut Visual v.2.15.0 on 2021-03-19

Figure S3. The output of the Alamut Visual version 2.15 (SOPHiA GENETICS, Lausanne, Switzerland) annotation tool when indicating a duplication of any one of the guanines in the Chr22(GRCh38:)g.50,721,505-50,721,512 region. Regardless of which G is selected in this homopolymer run as duplicated the variant is annotated as a duplication of the final G at the 3' end of the gene according to standard HGVS nomenclature (https://varnomen.hgvs.org/). The high frequency of the variant reported in ESP is likely to be artifactual since coverage at this position is low in the ESP database and the frequency is low in newer population databases like gnomAD. Moreover, even in gnomAD the variant is only present in the filtered reads, suggesting low variant quality.
